# Development of an Open-Access Action Observation Video Library for Upper Limb Motor Rehabilitation

**DOI:** 10.64898/2026.06.10.26355108

**Authors:** Madison Medina, Lewis A. Wheaton, Veronica Rowe

**Affiliations:** Department of Occupational Therapy, Byrdine F. Lewis College of Nursing and Health Professions, Georgia State University; School of Biological Sciences, Georgia Institute of Technology

**Author notes:** Corresponding Author: Veronica T. Rowe, MS(R), PhD, OTR/L, CBIST, FNAP, Department of Occupational Therapy, Georgia State University Urban Life Building, 140 Decatur St., Suite 1242, P.O. Box 3995 Atlanta, GA 30302-3995, Office.

## Abstract

**Background:** Occupational therapists can improve stroke survivors’ hand and arm movement and participation in daily activities through action observation (AO). AO involves watching another person’s hand or arm complete a movement or task. While research generally supports the use of AO with stroke survivors, there are limited AO videos are available to occupational therapists which makes applying AO challenging.

**Objective:** The purpose of this work is to develop structured and widely accessible tool to support access to AO for stroke survivors, occupational therapists, and researchers.

**Methods:** To develop an AO video library for stroke rehabilitation, functional and non-functional upper limb task deficits were first identified through clinical observations and clinician interviews to establish a prioritized list of daily activities. In collaboration with media production specialists, healthy adult volunteers were recruited and filmed performing these tasks from both first-and third-person perspectives. The recorded videos were then systematically edited, enhanced with instructional title slides, and distributed via a public YouTube channel for clinical application and a categorized digital repository for research purposes.

**Results:** Initial assessments revealed a complete lack of familiarity, awareness, and utilization of AO resources among local occupational therapists, despite high perceived clinical utility. To address this gap, a final library of 150 tasks was established, resulting in the production of 419 finalized, standardized videos featuring six healthy volunteers. For clinical application, these videos were hosted on a free, public YouTube channel organized into 18 functional playlists, while a parallel set was structured into distinct movement categories for research repository storage.

**Conclusion:** By providing a structured and highly accessible tool, this repository enables clinicians, researchers, and caregivers to readily implement evidence-based action observation interventions in both clinical and home settings.

## Introduction

Each year in the United States, over 795,000 individuals have a stroke, most of which are first or new strokes (Centers for Disease Control and Prevention [CDC], 2022). Following a cerebrovascular accident, stroke survivors often require caregiving because stroke is a primary cause of long-term disability (Mack & Hildebrand, 2023). Strokes can significantly impair an individual’s body structures and body functions which can affect a person’s motor control, sensation, and proprioception (Raghavan, 2015). Nearly 80% of stroke survivors have a motor impairment, with upper limb impairments leading to more functional deficits than lower limb impairments since upper limbs are involved in many daily activities (Pollock et al., 2014; Raghavan, 2015). Also, upper limb impairments can result in learned nonuse, which is when a stroke survivor learns to stop using their affected upper limb because it is easier to use their unaffected upper limb (Raghavan, 2015). Limited use of the affected upper limb can contribute to soft tissue changes and complications such as pain and contractures (Raghavan, 2015). It is important to address upper limb impairments in stroke rehabilitation, given they can lead to physiological changes and hinder participation in daily activities (Raghavan, 2015; Pollock et al., 2014).

Action observation (AO) is one relatively new intervention used by occupational therapists for hand and upper extremity rehabilitation to facilitate motor recovery for stroke survivors (Buchignani et al., 2019; Braun et al., 2013; Borges et al., 2022). AO involves viewing video recordings or in-person demonstrations of meaningful, goal-directed actions to stimulate the mirror neuron system (MNS; Kim et al., 2017; Buchignani et al., 2019). During AO, clients are instructed to closely watch the actions of healthy individuals and then attempt to replicate those actions to the best of their abilities (Peng et al., 2019; Zhang et al., 2023). Actively practicing the actions shown in AO videos is known as action execution (AE).

The discovery of AO began in the 1990s when researchers studying monkeys discovered a group of neurons in the premotor cortex that could be activated through both observation of a movement and motor execution of the same movement (Cengiz et al., 2018). This group of neurons was coined the MNS (Cengiz et al., 2018). Further research determined that humans also have an MNS that can be activated by observing movements, even in the absence of movement (Cengiz et al., 2018; Goldberg & Nilsen, 2020). The MNS has four key features: it is purposeful, context-dependent, experience-based, and multisensory (Shamili et al., 2022). The discovery of the MNS led to investigations in AO as an intervention to promote motor learning and enhance motor performance for various populations including individuals with stroke, cerebral palsy, Alzheimer’s disease, and Parkinson’s disease (Buchignani et al., 2019; Kim et al., 2017).

AO has been found to activate a bilateral premotor, parietal, and sensorimotor network in the brain which might facilitate movement execution and motor learning by facilitating the excitability of the motor system (Borges et al., 2022). Observing functional tasks in AO videos appears to stimulate the MNS more than simple actions because functional activities better reflect the four key features of the MNS (Shamili et al., 2022). Further, the effectiveness of AO is enhanced when the observer is able to hear the sounds of the actions being performed in the videos and when the tasks in the videos are broken into individual components, e.g., separating the task of drinking into multiple videos like pouring, reaching, and bringing the drink to mouth (Mancuso et al., 2021).

Stroke survivors with different levels of upper limb impairments can benefit from AO (Mancuso et al., 2021). AO is an appropriate intervention for stroke survivors because it promotes neuroplasticity by rearranging partially damaged motor circuits and does not require movement of a paretic upper limb (Borges et al., 2022; Franceschini et al., 2022; Craighero et al., 2023). Upper limb impairments after stroke can include long-lasting motor dysfunction and limited voluntary movement of the upper limb, and stroke survivors with these impairments must relearn motor skills or acquire new motor skills for improved upper limb performance (Zhang et al., 2019; Borges et al., 2022). By facilitating neuroplasticity through the activation of damaged motor circuits, AO can promote motor learning and recruit the motor system to improve stroke survivors’ upper limb function, even if they are unable to perform the observed tasks with their upper limbs (Yu & Park, 2022; Mancuso et al., 2021; Craighero et al., 2023).

Although AO can be beneficial for stroke survivors with varying degrees of impairment, the AE phase may be challenging for those with severe impairments who have little to no upper limb movement (Mancuso et al., 2021; Rungsirisilp et al., 2023). Some studies have used mental imagery (MI) with AO in place of AE (Rungsirisilp et al., 2023; Binks et al., 2023). MI is performed by mentally rehearsing an action, practicing an action in one’s mind without attempting to physically perform the action (Rungsirisilp et al., 2023). Using MI with AO involves simultaneously imagining oneself performing the AO tasks while watching the AO videos (Rungsirisilp et al., 2023; Binks et al., 2023). This allows stroke survivors with severe upper limb impairments to mentally practice the AO tasks rather than physically practice the AO tasks. Combining MI with AO can result in motor improvement, “cortical excitation of the affected sensorimotor hand region,” and improved upper limb function (Rungsirisilp et al., 2023, p. 4940). Moreover, using AO and MI together “activates motor execution-related brain regions more effectively than either technique alone” (Rungsirisilp et al., 2023, p. 4932).

While AO is utilized to promote neuroplasticity for stroke survivors with mild to severe impairments, there is currently mixed evidence regarding the effectiveness of AO for stroke survivors. The most recent Cochrane Review on AO revealed that this intervention can lead to improved upper limb motor function for stroke survivors (Borges et al., 2022). While AO was shown to have a large effect on hand function, AO had a small effect on arm function (Borges et al., 2022). The Cochrane Review also indicated that the certainty of current evidence was very low to low, leading to limited confidence in the effect estimates (Borges et al., 2022). At the time of publication for the Cochrane Review, four other systematic reviews on the effects of AO for stroke survivors had been published (Borges et al., 2022). Overall, the results of these systematic reviews aligned with the conclusions of the Cochrane Review, but Peng et al. (2019) found that the effect size of AO on arm function was larger than that of the Cochrane Review, and Ryan et al. (2021) found stronger evidence for AO compared to the Cochrane Review. Despite the few differences among these systematic reviews and the limited certainty of current evidence, these studies demonstrated that AO does not have adverse effects, is inexpensive, and can improve upper limb function, so it can be considered a useful occupational therapy intervention for stroke survivors (Borges et al., 2022).

Like the mixed evidence about the effects of AO on upper limb motor function, there is also conflicting evidence regarding the effects of AO on activities of daily living (ADLs). The latest Cochrane Review showed there was no effect of AO on ADL performance for stroke survivors compared to the control groups (Borges et al., 2022). In contrast, multiple other studies have found that AO may enhance engagement in various occupations. Yu & Park (2022) found AO can lead to improvements in ADL performance, with greater improvements when AO videos were shown from a first-person perspective. They reported significant differences in ADL performance before and after AO for personal hygiene, feeding, toilet transfers, and dressing (Yu & Park, 2022). Goldberg & Nilsen (2020) conducted a systematic review that also concluded that AO can significantly improve ADL performance. A randomized clinical trial by Shamili et al. (2022) found that using meaningful activities for AO can improve cortical excitation, actual task performance, and perceived performance and satisfaction of meaningful occupations. This is because using tasks and activities selected by and meaningful to stroke survivors likely leads to more intrinsic motivation and volition during AO sessions and improved occupational performance (Shamili et al., 2022). In a meta-analysis by Peng et al. (2019), AO had a moderate to large effect size on ADL outcomes, likely because the analyzed videos included goal-directed and functional activities. Additionally, Mancuso et al. (2021) reported AO resulted in clinically meaningful improvements in functional outcomes for stroke survivors which could lead to increased independence in daily activities. Due to the discrepancies among existing literature, more research is needed to better understand the impact of AO on occupational performance.

Several issues related to implementing AO into clinical practice are present in the literature. For instance, there is a lack of clear intervention protocols for AO, high heterogeneity in how AO is implemented with stroke survivors, and a lack of understanding about which clients are appropriate for this intervention (Franceschini et al., 2022; Ietswaart et al., 2015; Peng et al., 2019). There is also a need for further understanding of the mechanisms for this intervention (Zhang et al., 2019). If clinicians have conceptual confusion about AO and its methods and lack guidance on how to implement this into clinical practice, it can lead to underutilization of AO in clinical practice (Ietswaart et al., 2015). Clarity on the fundamentals and benefits of AO is needed to promote clinician utilization (Ietswaart et al., 2015).

To clarify the protocol for implementing AO in stroke rehabilitation, Zhang et al. (2023) recently conducted a systematic review addressing confusion surrounding its clinical application. By reviewing 29 studies, Zhang et al. (2023) provided valuable insights on how to implement AO with stroke survivors. They found that a majority of studies on AO for stroke survivors used short video lengths for the AO intervention, typically a few minutes long, and provided the stroke survivors with rest in between watching the videos. Also, stroke survivors who began AO more than a month after stroke onset had better outcomes and required fewer weeks of intervention to achieve significant differences in outcome measures than those who began the intervention less than one month after stroke onset. Consequently, Zhang et al. (2023) recommended starting AO at least 23 days after stroke onset. Further, they determined that AO should be performed for 30-40 minutes in a single session to account for stroke survivors’ limited attention and still achieve effective results. Finally, although Zhang et al. (2023) was unable to determine the optimal duration for implementing AO based on the literature, they suggested that stroke survivors perform AO at least three to five times a week for a minimum of four weeks.

Zhang et al. (2023) helped create a more defined protocol for AO for stroke survivors. However, their study was published less than a year ago, and it is likely that their findings have not yet been implemented in clinical settings due to overall delays in applying current evidence to stroke rehabilitation (Bayley et al., 2012; Juckett et al., 2020). In fact, Juckett et al. (2020) stated there is a 17-year lag in implementing research into clinical practice for healthcare settings. Therefore, based on current problems listed in the literature, including the lack of clarity for AO delivery and dosage, it appears there is a lack of knowledge translation (KT) for AO and hesitation to use AO in clinical practice (Ietswaart et al., 2015).

KT can be defined as “a dynamic and iterative process that includes the synthesis, dissemination, exchange and ethically sound application of knowledge to improve health, provide more effective health services and products, and strengthen the health care system” (Straus et al., 2009, p. 165). KT leads to the use of knowledge in clinical practice after knowledge is created through research studies and disseminated (Straus et al., 2009). KT targets stakeholders beyond just healthcare professionals, such as patients and policymakers (Straus et al., 2009). Two forms of KT are end-of-grant KT and integrated KT (Barwick et al., 2020). End-of-grant KT involves a one-way process where stakeholders have limited involvement in knowledge creation, while integrated KT involves a collaborative process between knowledge producers and stakeholders throughout all stages of knowledge creation and translation (Barwick et al., 2020). Integrated KT aims to create research that is more pertinent to stakeholders than end-of-grant KT (Canadian Institutes of Health Research, 2015). Effective KT is critical in healthcare because inadequate KT can lead to a lack of evidence-based practice causing healthcare professionals to provide suboptimal care and use ineffective methods (Straus et al., 2009; Graham et al., 2006).

Although research supports many occupational therapy interventions for stroke rehabilitation, including AO, translating research to occupational therapy practice continues to be a challenge (Walker et al., 2013). In general, occupational therapists do not fully incorporate recent research into intervention sessions for stroke survivors (Bayley et al., 2012). One barrier to evidence-based practice is a lack of time for occupational therapists to find and read articles and determine how to implement research into clinical practice (Lindström & Bernhardsson, 2018). Another obstacle is accessibility to resources such as scholarly magazines, journals, and databases (Lindström & Bernhardsson, 2018). Some occupational therapists have poor confidence in their research skills or lack research skills, leading to difficulty understanding and implementing research into intervention sessions. Also, occupational therapists have difficulty implementing evidence-based practices due to the costs of interventions, logistical challenges, and insufficient access to equipment and resources (Juckett et al., 2020).

A delay in KT limits patient outcomes and hinders evidence-based practice (Bayley et al., 2012). The delay between research dissemination and implementation of research into clinical practice for stroke rehabilitation needs to be reduced, especially as the number of individuals with stroke increases. Therefore, this project aims to answer the following PICO question: “To what extent can KT (I) increase the utilization of AO for hand and upper extremity rehabilitation (O) for stroke survivors (P)?” The purpose of this study was to facilitate KT for AO to increase the use of this intervention during occupational therapy sessions with stroke survivors in an outpatient clinic and future research endeavors. The focus of this project is to create AO videos and gather educational resources about AO to deliver to occupational therapy practitioners and researchers.

## Methods

An application for designation of not human subjects research for this project was submitted to the Georgia State University Institutional Review Board (IRB). IRB approval was granted, and the submission was determined to be exempt from review. The overall development of the videos and instructional materials involved an iterative design process as summarized in Figure 1.

**Figure 1.**
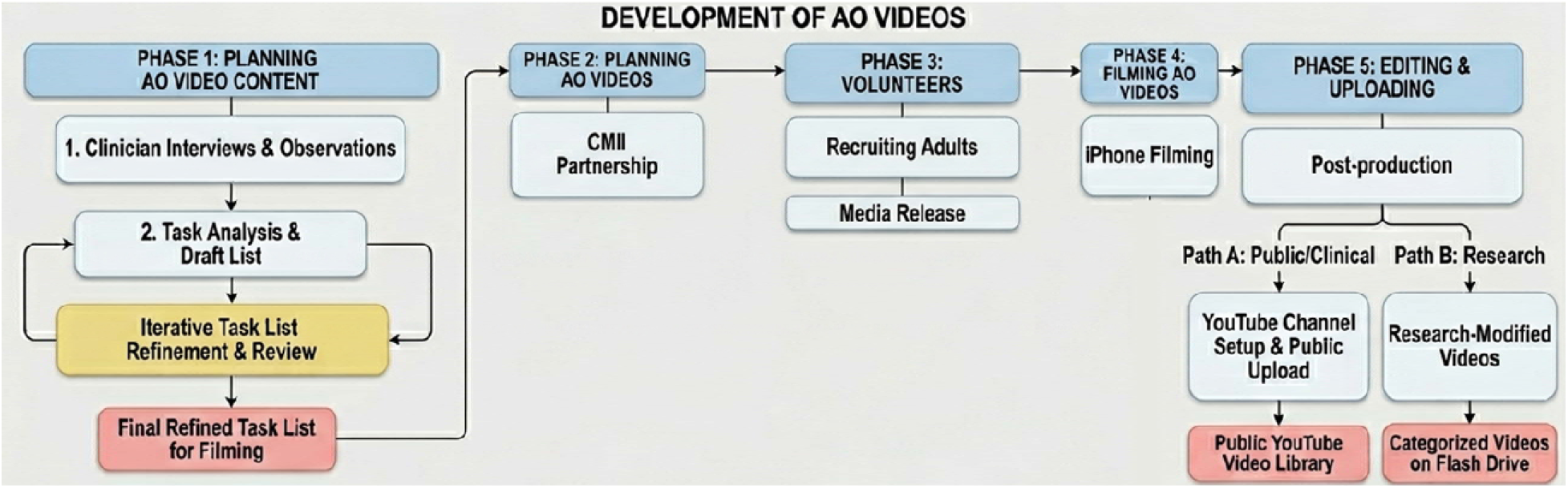
Design process of AO Videos.

### Development of AO Videos

#### Phase 1: Planning AO Video Content

To inform the content for the AO videos, we first evaluated the present state of AO was in clinical practice. Observations and informal interviews with occupational therapists occurred at Encompass Health Rehabilitation Hospital, Emory Johns Creek Hospital, Children’s Healthcare of Atlanta, and the Shepherd Center, all located in the metropolitan Atlanta, GA community. No AO resources were discovered at these sites. However, while observing clients at these sites, we gained insight into clients’ deficits and functional goals which helped establish the basis for the AO content.

Next, observations were completed at Rehabilitation Without Walls (RWW; Lawrenceville, GA) to determine the types of tasks that would be appropriate for AO videos for stroke survivors receiving occupational therapy services in an outpatient setting. At RWW, a variety of functional deficits were observed including challenges with folding laundry, tying shoes, pulling up pants, feeding oneself with a spoon, writing personal information with good legibility and letter placement, navigating a cell phone to make calls or send text messages, opening food packaging, cutting with scissors, and putting on a zip-up jacket. These clinician interviews helped identify the list of proposed activities for AO video content.

An initial list of AO tasks was formed based on the observations of stroke survivors and discussions of common functional challenges experienced by stroke survivors at RWW. During this initial AO task ideation, a task analysis of typical daily activities performed by stroke survivors. The task analysis broke down daily activities, like feeding, grooming, and meal preparation, into their task components to determine which components should be included in AO videos. Further, existing AO videos on YouTube to identify if any of these task components were already readily available as AO videos or if these tasks were missing from the current library of AO videos on YouTube.

The initial draft was reviewed by all members of the research team. The list of AO tasks was developed to include non-functional finalized actions and non-finalized actions since the list previously primarily contained functional finalized actions. Additionally, the AO task list was revised to remove any FTHUE items that only required isometric contraction of the upper limb since these tasks didn’t require active movement of the arm. After the list of AO tasks was updated based on initial feedback from my site mentors, the list was reviewed again by both mentors. At this stage in the development, the list contained over 200 tasks. To create a more concise list of AO tasks, the site mentors were consulted to remove less relevant and low-priority tasks.

Next, the tasks on the list were categorized. This ensured a sufficient number of tasks were included in each task category and to verify if there were enough appropriate tasks for research and clinical applications. The number and types of tasks in the categories were not disproportionate. Hence, the tasks listed in these categories did not require further modification and were selected to be filmed for the AO videos.

#### Phase 2: Planning AO Videos

The Creative Media Industries Institute (CMII) is a content creation center at Georgia State University and has several production and post-production studios. Students and faculty from the CMII have expertise in filming and editing for film and TV production and have access to high-quality technology. Therefore, a student and faculty member from the CMII were chosen to partner with this project to assist with planning and filming the AO videos.

#### Phase 3: Recruiting Volunteers to Feature in the AO Videos

The individuals featured in the AO videos needed to be healthy volunteers with no upper limb impairments. The inclusion criteria for volunteers for the AO videos were adults between 18 - 89 years old who had no upper limb impairments to either arm, did not use upper limb prosthetics, could read and write in English, and could understand spoken English. Volunteers were excluded if they were under 18 years old, over 89 years old, unable to follow instructions in English due to cognitive abilities or language barriers, had an amputated upper limb, or had limited upper limb motor function (e.g., hemiplegic arm).

Convenience sampling was utilized to recruit volunteers. Those meeting the inclusion criteria were contacted electronically via text message and asked if they were interested in volunteering for the AO videos. Volunteers who agreed to participate were asked to sign a media release form, giving their approval to use videos with their images for educational and research purposes. The media release form was verbally reviewed with volunteers and any questions were answered. Voluntary consent was obtained with the volunteers’ signatures on the hard copy media release form. There was no compensation for volunteers. The costs to the volunteers that may have resulted from participation in the project included transportation costs for commuting to film sites. There was no direct benefit to the volunteers, however, participants were informed that these videos will benefit occupational therapists, researchers, and stroke survivors. The risk of this study was equivalent to that of engaging in everyday activities, however, this was minimal. In order to minimize this risk, volunteers could have terminated their participation in filming at any point.

#### Phase 4: Filming AO Videos

All filming was completed on an iPhone 15 Plus. Two volunteers were filmed at Georgia State University. The first filming session lasted four hours. A variety of third-and first-person videos were filmed during this session. The remaining videos were filmed off campus.

#### Phase 5: Editing and Uploading AO Videos

All video files were shared from the iPhone to a MacBook Air using AirDrop, and the files were uploaded to iMovie for editing. Each video was edited individually. Examples of edits made to the videos using iMovie included trimming videos, cropping videos, altering the brightness, flipping videos horizontally, rotating videos clockwise, copying and pasting sections of the videos, modifying video speed (i.e., slow speed by 15-50%), removing background noise, reversing videos, stabilizing shaky videos, adding fade to black transitions, adding freeze frames, removing audio from a video, and adding text to the videos.

Title slides for all videos were created using Canva. A screenshot from each video was uploaded to Canva, and for most of the screenshots, the background of the image was removed using Canva’s background remover tool or through manual editing. The images were then positioned next to text that described the primary action of the video. The final title slides were downloaded as JPEGs, uploaded to iMovie, and inserted at the start of each corresponding video on iMovie. An end screen slide was also designed on Canva and uploaded to iMovie. The end screen was the same for all videos and was added at the end of each video. The length of the end screen was set to at least five seconds for each video because this was a required end screen length from YouTube. Videos were organized and uploaded based on clinical or research use.

Clinical use: A YouTube channel was created to compile all final AO videos into a video library for clinical use. A logo and a banner were designed on Canva and uploaded to the YouTube channel. The edited videos were uploaded to the YouTube channel. The videos were given a YouTube video title, and time stamps were added to the description of the videos. Videos were assigned to a playlist on the YouTube channel, the intended audience was set in the video settings, the title slide JPEGs were uploaded as thumbnails, and the visibility of the videos was set to public. Then, end screen templates were created on the YouTube channel. The end screen template was selected and uploaded separately for every video.

Research use: For research purposes, the AO videos that were uploaded to the YouTube channel were modified on iMovie to remove the end screen. These modified videos were then saved, and the files were named based on the tasks shown in the videos. The files were uploaded to a flash drive. On the flash drive, the videos were categorized into 4 folders: functional finalized actions, non-functional finalized actions, non-finalized actions, and FTHUE.

## Results

### Current AO Resources

When observing occupational therapists at the four sites in metro-Atlanta, the informal discussions revealed that these clinicians are not currently implementing any AO resources into their treatment sessions. Additionally, they were not aware of any AO resources available at their sites. Moreover, all these occupational therapists were unfamiliar with AO and had not previously learned about AO. After a brief description of AO to the clinicians, most of them asked follow-up questions to better understand AO and asked how to access AO resources. Some of these occupational therapists responded by stating AO would be beneficial for their current clients which demonstrates occupational therapists perceive AO resources would be useful in various rehabilitation settings.

### List of AO Tasks

A final list of 150 tasks was included for AO tasks. Most of the AO tasks were seated to promote safe execition for stroke survivors who have diminished coordination and balance.

Fifteen (15) of the 17 FTHUE items were included in the list of AO tasks. The FTHUE items excluded from the AO task list were “associated reactions” and “hold a pouch” since these items required a static upper limb position. The 150 AO tasks were grouped into different categories for researchers and clinicians. For research purposes, AO tasks were categorized into functional finalized actions, non-functional finalized actions, and non-finalized actions. For clinical purposes, AO tasks were placed into 17 categories that best described the primary actions of the videos (see Supplemental A). All tasks required no to minimal supplies, and those that required supplies used common household items. The AO tasks intentionally used easily accessible and inexpensive supplies to minimize a potential barrier to completing AO and AE at home.

**Figure 2.**
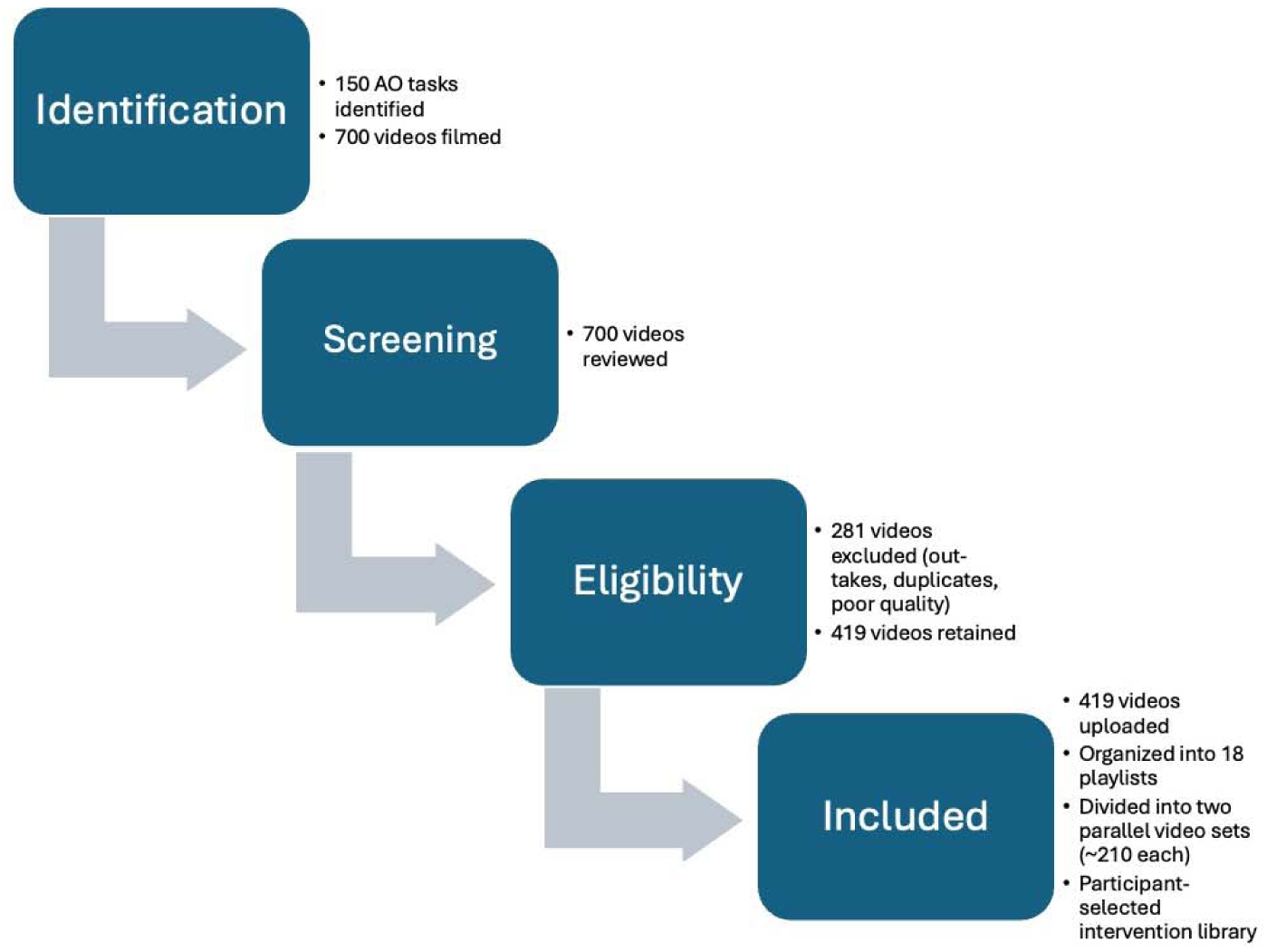
Development of the Action Observation Video Library. Approximately 150 action observation tasks were identified and used to guide the filming of approximately 700 videos. Following review, editing, and upload, 419 videos were retained. The final library was organized into two parallel sets of approximately 210 videos each and categorized into 18 functional playlists to support graded action observation intervention delivery.

### Volunteers

Six individuals volunteered to be featured in the AO videos. The volunteers included three white females, one black female, and two white males. All volunteers were between 23 and 31 years old and right-handed. Volunteers were asked to wear solid-colored clothing and remove accessories including watches and jewelry. For all filming sessions, volunteers were instructed to complete each task at a slightly slow pace to make their hand and arm movements easier for stroke survivors to watch. The volunteers primarily used their dominant arm for unilateral tasks. For activities that required repetitions of a movement or task, such as range of motion movements and opening and closing containers, volunteers completed between three to five reps of the movements. A small number of repetitions was used for AO tasks to reduce overall filming time for volunteers. For many of the functional tasks, such as brushing teeth or folding clothes, volunteers typically performed the tasks for 20 to 40 seconds.

### AO YouTube Channel

A YouTube channel named *ActionObservationOT* was developed to host the final AO videos for clinical use. The AO video library was uploaded to YouTube for occupational therapists, clients, and caregivers because it is a free and widely available platform. A description of the YouTube channel was added to help users understand the purpose of the channel.

Approximately 700 videos were filmed of the volunteers. Each of the videos was reviewed and outtakes were deleted. A total of 419 AO videos were edited on iMovie and uploaded to the YouTube channel. Only a few AO videos demonstrating FTHUE items were uploaded to the YouTube channel because most of the FTHUE items were less relevant for clinical purposes. Each video was given a title that described the task in the video and a thumbnail showing a screenshot of the task (Figure 3). The AO videos were set to public visibility to make the YouTube channel easy to navigate, especially for individuals who have limited experience using YouTube.

**Figure 3:**
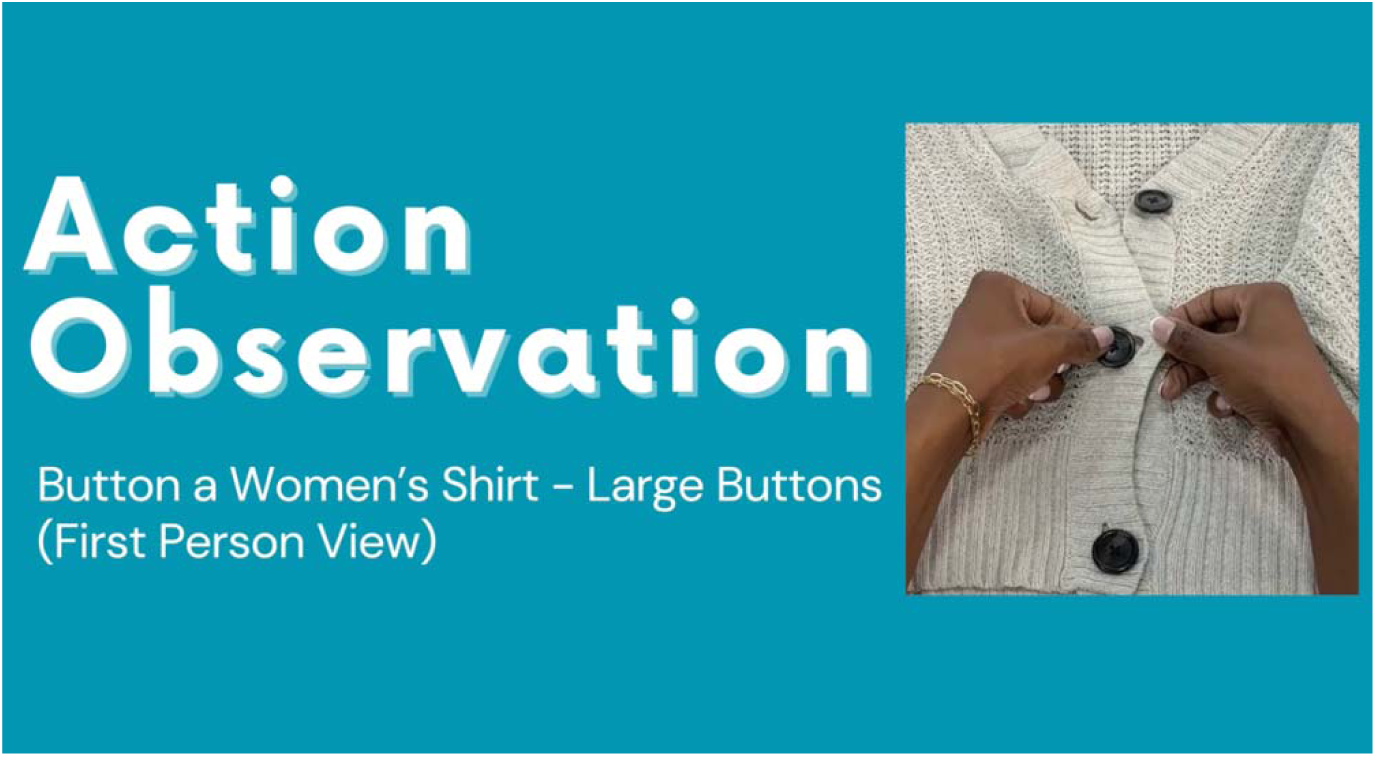
Example of an AO Video Thumbnail

All the videos were categorized by areas of function and assigned to one of 18 available playlists (see Figure 4). Examples of playlists were fine motor activities, kitchen tasks, upper body dressing, and laundry tasks. The purpose of the playlists was to improve the organization of the channel to allow stroke survivors and their caregivers to easily find AO videos that are most relevant to their goals. Further, when a playlist is selected on YouTube, it will automatically play other videos from that playlist. This will reduce the amount of time users spend searching for similar AO videos. To make the playlists easily accessible, a clickable link to the related playlist was included at the end screen of each AO video. Using playlists to group the AO videos aimed to enhance the usability of the YouTube channel.

**Figure 4:**
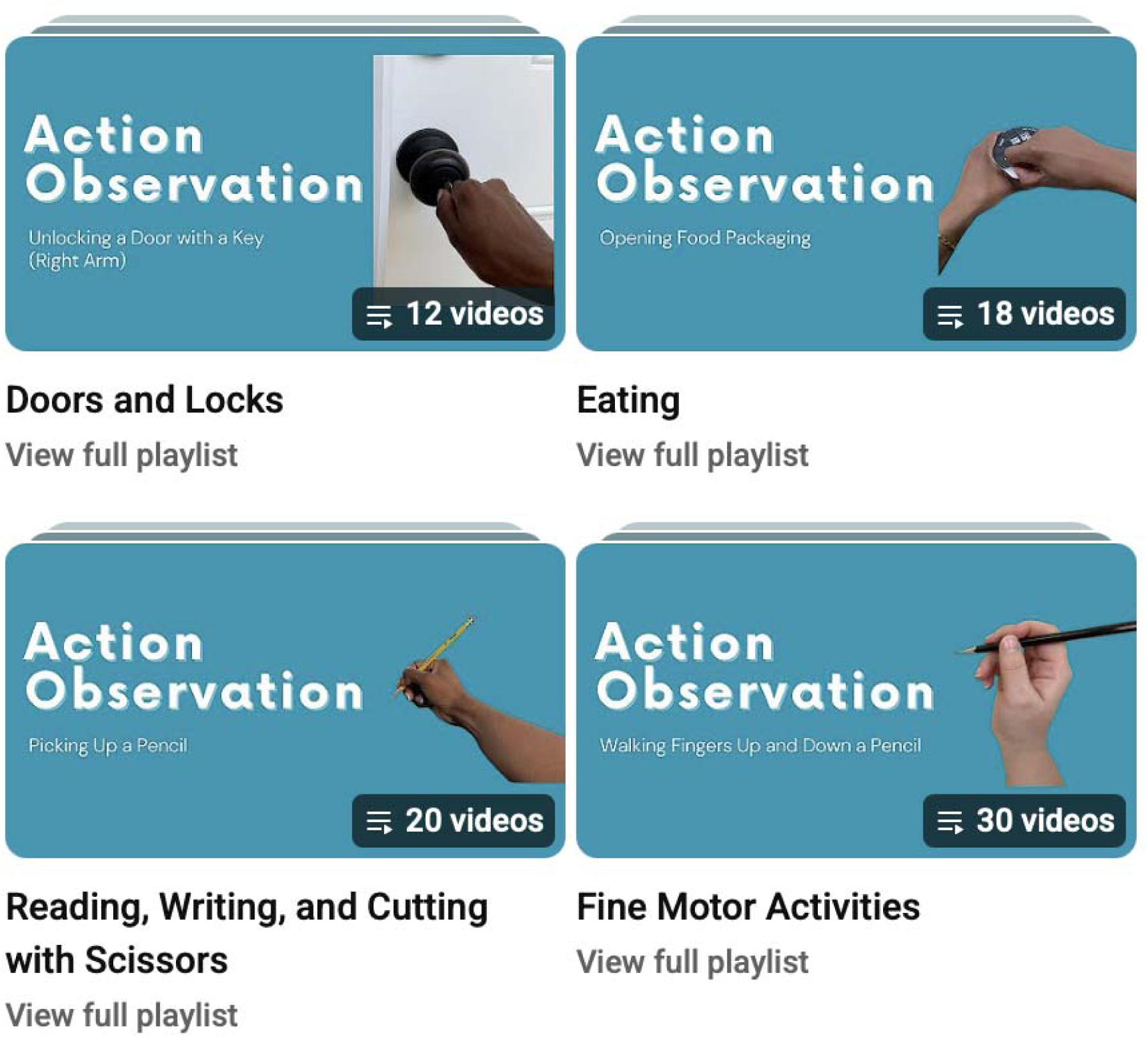
YouTube playlists organized by actions

Finally, the videos that were uploaded to YouTube were modified in iMovie. Then, these modified videos were uploaded to flash drive folders and categorized into functional finalized actions, non-functional finalized actions, non-finalized actions, and FTHUE. Video files were also digitally stored on a cloud server and flask drive as needed for their future research.

## Discussion

Gaps in KT for AO and underutilization of AO were identified in AO literature and clinical practice. In general, there appears to be a lack of awareness about AO among occupational therapists in a variety of settings. Further, applying AO in stroke rehabilitation is currently challenging due to a variety of barriers, such as unclear implementation protocols and limited AO resources. Through this study, we were able to develop a clinician-informed video set that can serve as the basis for future studies and evaluation of AO.

Increasing awareness of AO among occupational therapists working in stroke rehabilitation will promote evidence-based practice which can lead to improved client outcomes. Also, having easier access to AO videos and educational resources about AO will increase the likelihood that AO will be implemented with stroke survivors. Increased utilization of AO with stroke survivors can improve their upper limb motor function and independence in ADLs, ultimately reducing caregiver burden and improving the quality of life of stroke survivors and their caregivers. Moreover, since a variety of AO videos are now available to researchers, they can use the videos to conduct additional research on AO. Future research can develop clearer intervention protocols for AO and further investigate the mechanisms of AO. These findings would assist occupational therapists in most effectively applying AO with stroke survivors, maximizing stroke survivors’ upper limb motor recovery and enhancing their occupational performance.

## Future Directions

Future projects can expand the efforts for engaging therapists in AO started in this project to other sites. Occupational therapists working in inpatient, outpatient, and home health settings would likely benefit from education about AO and access to AO resources tailored to their needs. Furthermore, more AO videos can be created and feature individuals of various ages, races, and hand dominance. This would increase the diversity of AO videos and allow stroke survivors to be better matched to the individuals featured in the AO videos. Finally, this project can be expanded to increase AO education and resources for other medical conditions such as Parkinson’s disease, cerebral palsy, or Alzheimer’s disease.

## Limitations

The AO videos developed for this project are limited because only young adults of two races were represented. A lack of diversity in the AO videos may increase the difficulty of matching stroke survivors to the AO videos and limit the outcomes of AO and AE (Robinson-Bert, 2023). Also, only right-handed individuals were featured in the videos which may make the videos less relevant to left-handed stroke survivors. Future AO videos should feature people of more races and ages and include left-handed individuals.

Also, limitations were present in the number of variations in task performance for the AO tasks. Most videos only showed one way of performing a task, but the way a task was performed on the YouTube channel is probably not how all stroke survivors perform that task. More variations in task performance would allow stroke survivors to choose the task performance that is most similar to their performance.

Additionally, while research supports AO tasks being filmed from a first-person perspective, some tasks for this project were only filmed from a third-person perspective due to logistical challenges. When attempting to film some tasks from a first-person perspective, volunteers’ hands and arms moved out of the video frame because the camera was close to their bodies. More guidance for filming from a first-person perspective would be helpful for future AO video development.

Lastly, since these videos were primarily filmed in public spaces, including a university and church, the noise in these environments could not be controlled. Some of the videos included significant background noise from people in the environment, and consequently, the audio was removed from these videos during the editing process. Ideally, the audio would have been included in all of the AO videos to increase the effectiveness of AO (Mancuso et al., 2021).

## Conclusion

Research on AO reveals this intervention can promote neuroplasticity for stroke survivors and lead to improvements in upper limb function and ADL performance. While there is sufficient evidence supporting AO for stroke rehabilitation, implementing AO into clinical practice has been limited due to a lack of AO resources and poor awareness of AO. In this work, AO videos of simple movements and daily tasks were developed by collaborating with occupational therapists working in stroke rehabilitation and researchers with expertise in AO. Over 400 AO videos were edited and uploaded to a public YouTube channel for use in research and clinical settings. These AO videos were also delivered to researchers to be used in future research on AO, including implementation studies, standardization assessments, and mechanistic studies of AO in clinical and non-clinical research.

Videos are available at: http://www.youtube.com/@ActionObservationOT

## Data Availability

All data produced in the present study are available upon reasonable request to the authors

## Supplemental A: Clinical and Research AO Video Categories

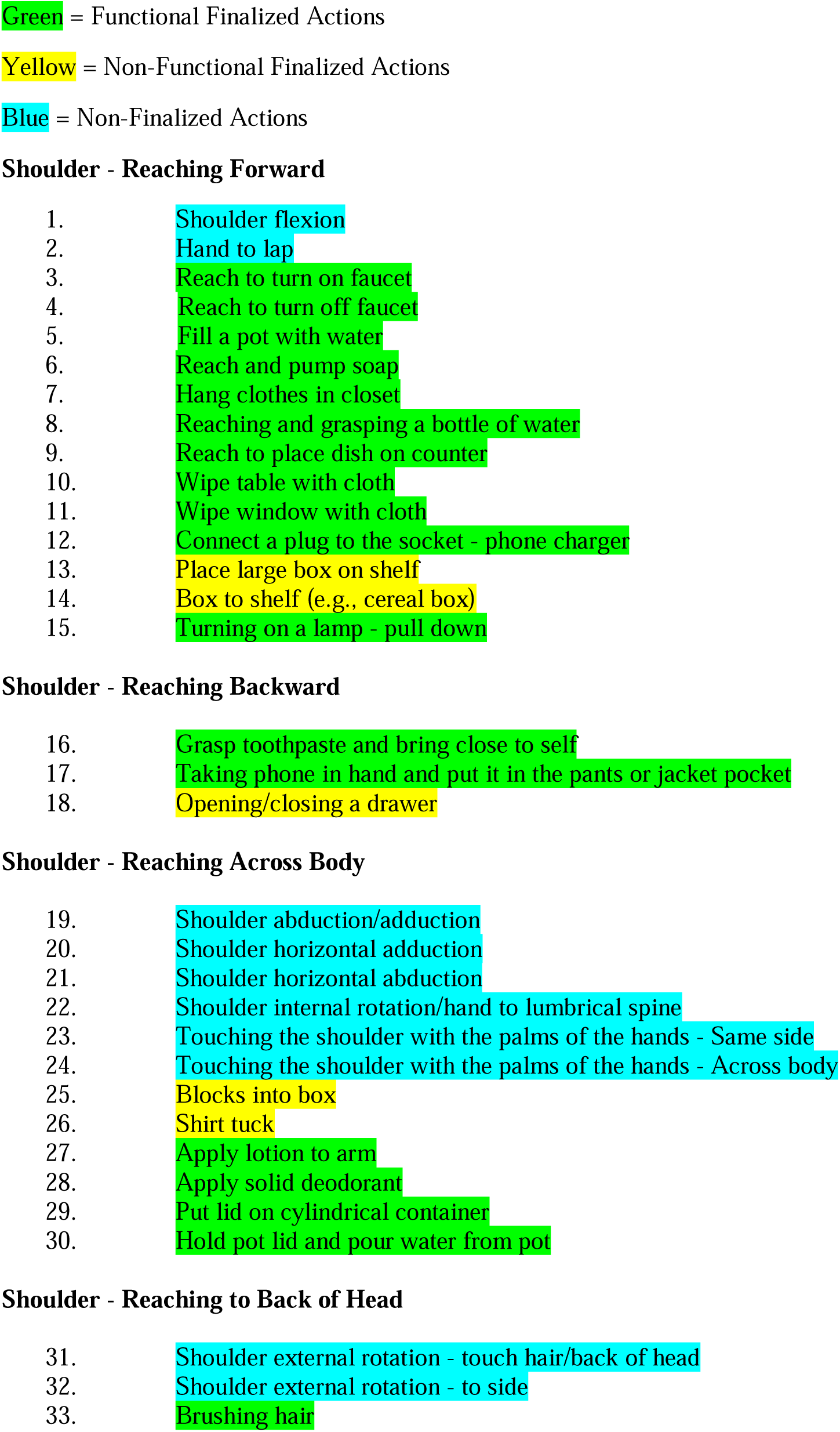
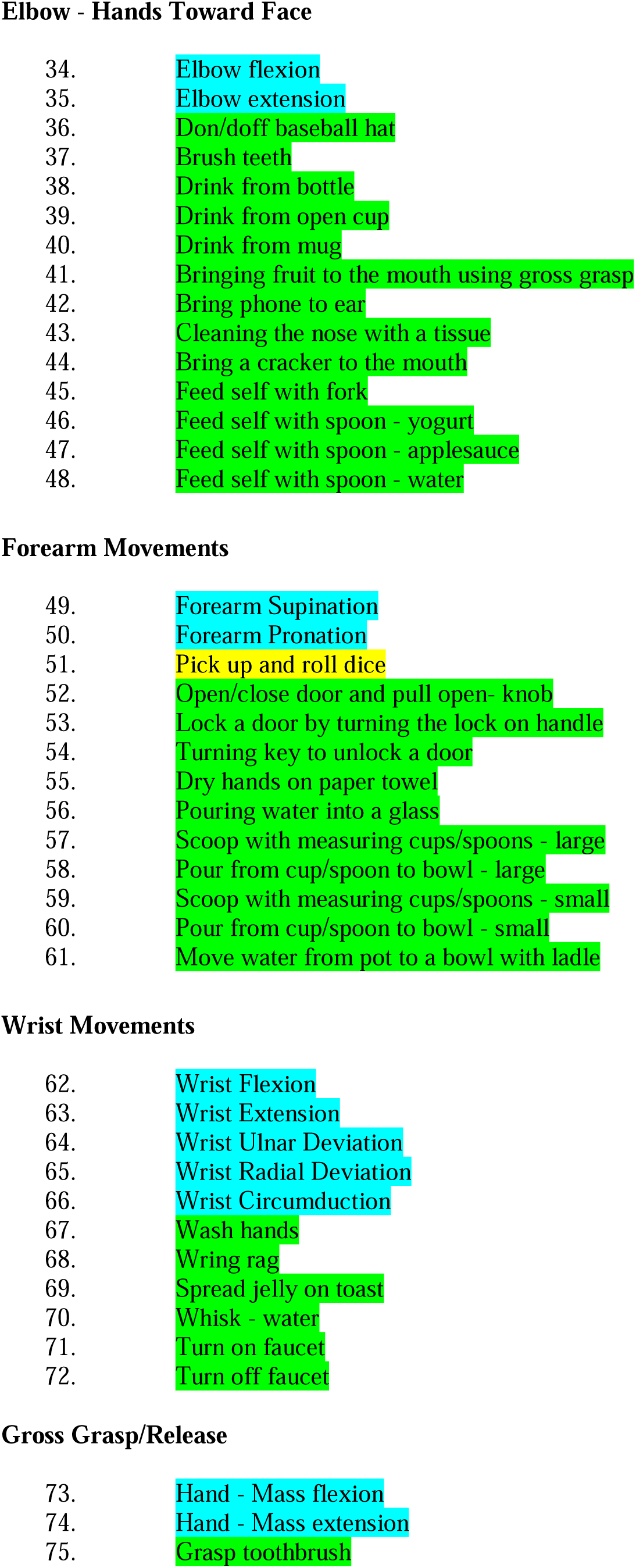
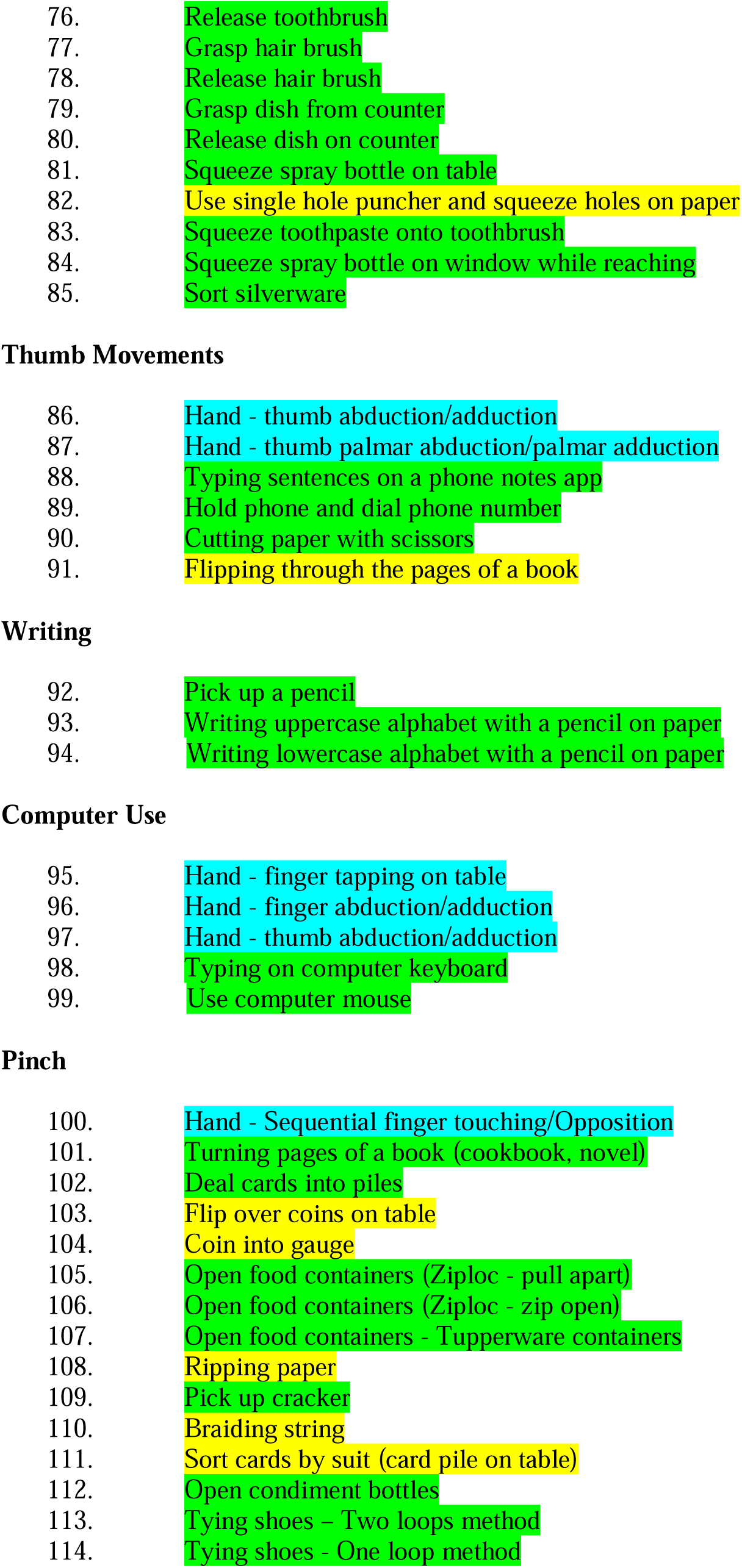
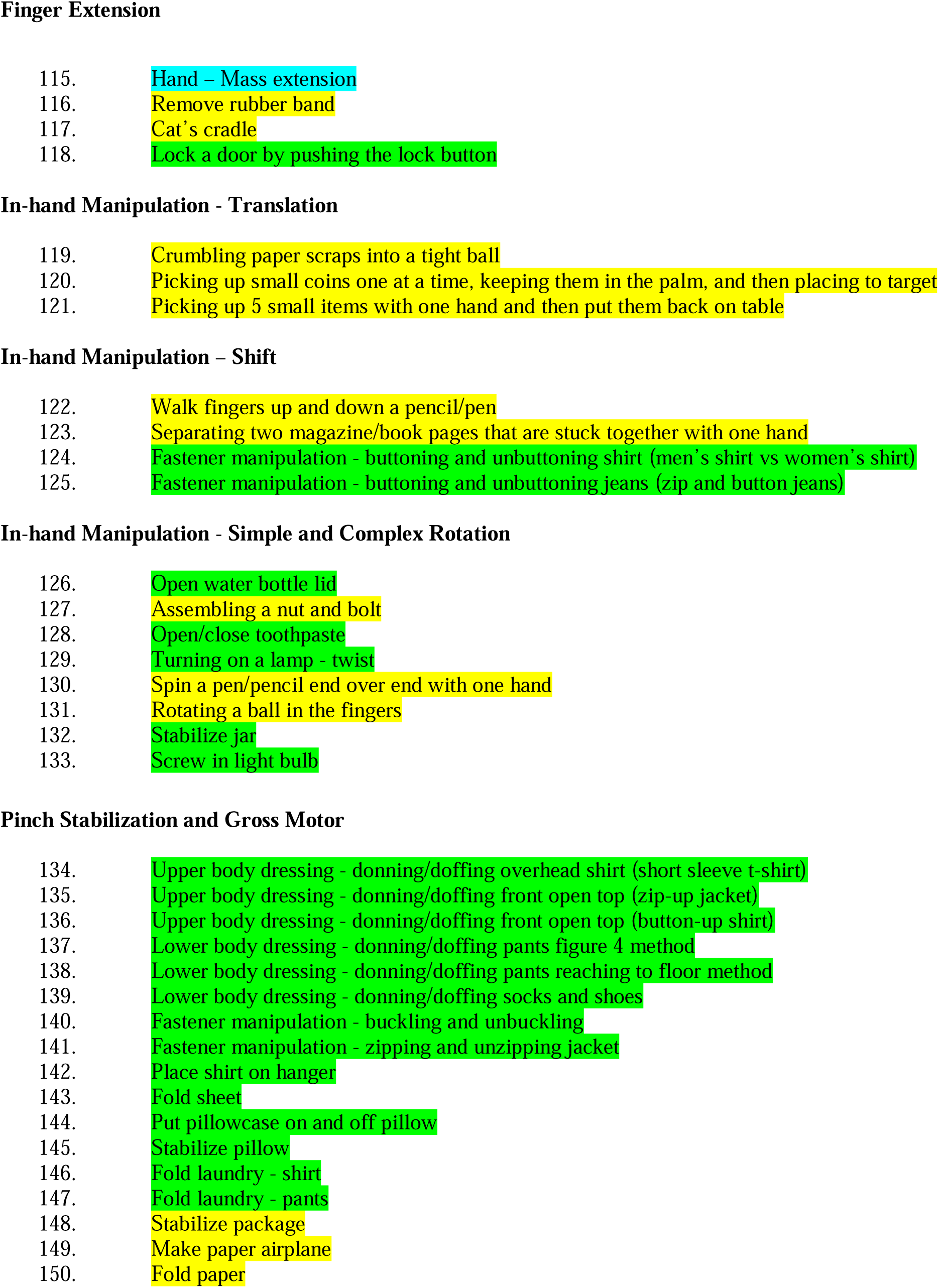

